# A handheld point-of-care system for rapid detection of SARS-CoV-2 in under 20 minutes

**DOI:** 10.1101/2020.06.29.20142349

**Authors:** Jesus Rodriguez-Manzano, Kenny Malpartida-Cardenas, Nicolas Moser, Ivana Pennisi, Matthew Cavuto, Luca Miglietta, Ahmad Moniri, Rebecca Penn, Giovanni Satta, Paul Randell, Frances Davies, Frances Bolt, Wendy Barclay, Alison Holmes, Pantelis Georgiou

## Abstract

The COVID-19 pandemic is a global health emergency characterized by the high rate of transmission and ongoing increase of cases globally. Rapid point-of-care (PoC) diagnostics to detect the causative virus, SARS-CoV-2, are urgently needed to identify and isolate patients, contain its spread and guide clinical management. In this work, we report the development of a rapid PoC diagnostic test (< 20 min) based on reverse transcriptase loop-mediated isothermal amplification (RT-LAMP) and semiconductor technology for the detection of SARS-CoV-2 from extracted RNA samples. The developed LAMP assay was tested on a real-time benchtop instrument (RT-qLAMP) showing a lower limit of detection of 10 RNA copies per reaction. It was validated against 183 clinical samples including 127 positive samples (screened by the CDC RT-qPCR assay). Results showed 90.55% sensitivity and 100% specificity when compared to RT-qPCR and average positive detection times of 15.45 ± 4.43 min. For validating the incorporation of the RT-LAMP assay onto our PoC platform (RT-eLAMP), a subset of samples was tested (n=40), showing average detection times of 12.89 ± 2.59 min for positive samples (n=34), demonstrating a comparable performance to a benchtop commercial instrument. Paired with a smartphone for results visualization and geo-localization, this portable diagnostic platform with secure cloud connectivity will enable real-time case identification and epidemiological surveillance.

**One Sentence Summary:** We demonstrate isothermal detection of SARS-CoV-2 in under 20 minutes from extracted RNA samples with a handheld Lab-on-Chip platform.

## Introduction

A novel coronavirus first identified in Wuhan (China) in December 2019 resulted in the implementation of stringent measures such as quarantine or air traffic closure as an effort to limit its spread. Declared as a pandemic by the World Health Organization (WHO), SARS-CoV-2 has spread globally with over 7.8 million cases and 431,541 deaths worldwide *(1)*. The ability to accurately and reliably diagnose SARS-CoV-2, causing COVID-19 disease, is crucial for containing the pandemic enabling a proper response at national and global scales.

This highly contagious virus belongs to the genus of *Betacoronavirus* of the *Coronaviridae* family *(2, 3)*. As with all the human coronaviruses (hCoVs) identified to date, SARS-CoV-2 also have zoonotic origins *(4)*. According to the WHO, the most common symptoms of this lower respiratory tract infection include fever, cough, fatigue and pneumonia which can severely develop into acute respiratory distress syndrome and hyperinflammation *(5)*. SARS-CoV-2 is characterized by presenting high transmissibility (mainly through aerosolized droplets from saliva or nose), high infectivity and long incubation period (from 0 to 14 days) *(2, 5)*. In particular, the high number of asymptomatic subjects (mainly children and young adults) might be one of the major contributions to the rapid spread of this disease. Currently, several drugs and vaccine candidates are being assessed in clinical trials but there is no effective antiviral treatment or vaccine available yet. Therefore, rapid diagnostics are essential to respond to the pandemic and reduce the spread of COVID-19 by identifying and isolating both symptomatic or asymptomatic patients *(2, 6)*. Notably, rapid and point-of-care (PoC) diagnostics are essential to expand, scale up and increase the accessibility of testing around the world.

Different methods have been proposed for the diagnosis of SARS-CoV-2 including antigen detection, serologic tests, computerized tomography (CT) imaging and nucleic acid amplification tests (NAATs) *(7, 8)*. Serologic and antigen-based detection tests commonly suffer from low sensitivity and specificity, being qualitative and only capable of diagnosing past infections. Currently, reverse transcriptase polymerase chain reaction using real-time benchtop platform (RT-qPCR) is considered the gold standard for COVID-19 diagnosis due to its capability to detect the presence of SARS-CoV-2 RNA close to the onset of symptomatic illness which is critical for isolation. Two RT-qPCR kits have stood out as a reference to most studies, respectively developed by the CDC and the Institut Pasteur (with WHO approval) *(7, 9)*. While these kits are restricted to equipped health centers and are limited by procurement delays, isothermal methods such as reverse transcriptase recombinase polymerase amplification (RT-RPA) or loop-mediated isothermal amplification (RT-LAMP) are an alternative that do not require thermal cycling and provide results within an hour *(10–20)*.

These methods are ideal for PoC applications as they (i) reduce turnaround time from sample-to-results, (ii) reduce hardware complexity required and (iii) provide high sensitivity and specificity *(21, 22)*. To date, reported isothermal assays for COVID-19 detection are based on colorimetric detection which relies on trained personnel and lab-based equipment for experimental performance *(10–12, 16)*. This represents a barrier to the implementation of portable devices for SARS-CoV-2 PoC diagnostics, which can be overcome by using label-free electrochemical-based DNA detection. Recently, in *Rodriguez-Manzano et al. (23)*, we reported an embedded Lab-on-Chip (LoC) device for label-free biosensing applications which consists of ion-sensitive field-effect transistors (ISFETs) manufactured in unmodified complementary metal-oxide-semiconductor (CMOS) technology *(24)*. This device has integrated thermal management and is capable of nucleic acid detection by monitoring pH changes that occur during nucleic acid amplification which are compatible with isothermal assays. The platform demonstrates versatility to a wide range of targets and is compatible with real-time RT-LAMP (RT-eLAMP), and sample types when coupled with a sample preparation module. Additionally, its use of standard electronic components promotes scalability and portability which ideally match the requirements of portable diagnostics.

In this work, we designed and optimized a LAMP assay targeting the *nucleocapsid* (N) gene of SARS-CoV-2 based on reported sequences *(25)*. To validate the assay we used a real-time benchtop instrument (RT-qLAMP) and screened 183 clinical samples obtained from nasopharyngeal, throat and nose swabs of symptomatic patients. The results were compared with RT-qPCR *(9)*, which was used as a gold standard. The RT-qLAMP assay showed a sensitivity of 90.55% and specificity of 100% with a limit of detection of 10 copies per reaction and average positive detection times of 15.45 ± 4.43 min. Furthermore, we implemented the developed assay on our ISFET-based LoC platform for rapid PoC detection (Fig. 1) screening 40 samples, including positive (n=34) and negative (n=6) samples. Results obtained by both instruments were comparable (p-value > 0.05), demonstrating the capability of our handheld diagnostic platform paired with a smartphone application to rapidly identify and quantify the presence of SARS-CoV-2 RNA with high reliability. This constitutes the first handheld molecular diagnostic for SARS-CoV-2 detection within 20 min from extracted RNA. Portable diagnostics with secure cloud connectivity will enable real-time case identification and epidemiological surveillance.

**Fig. 1:**
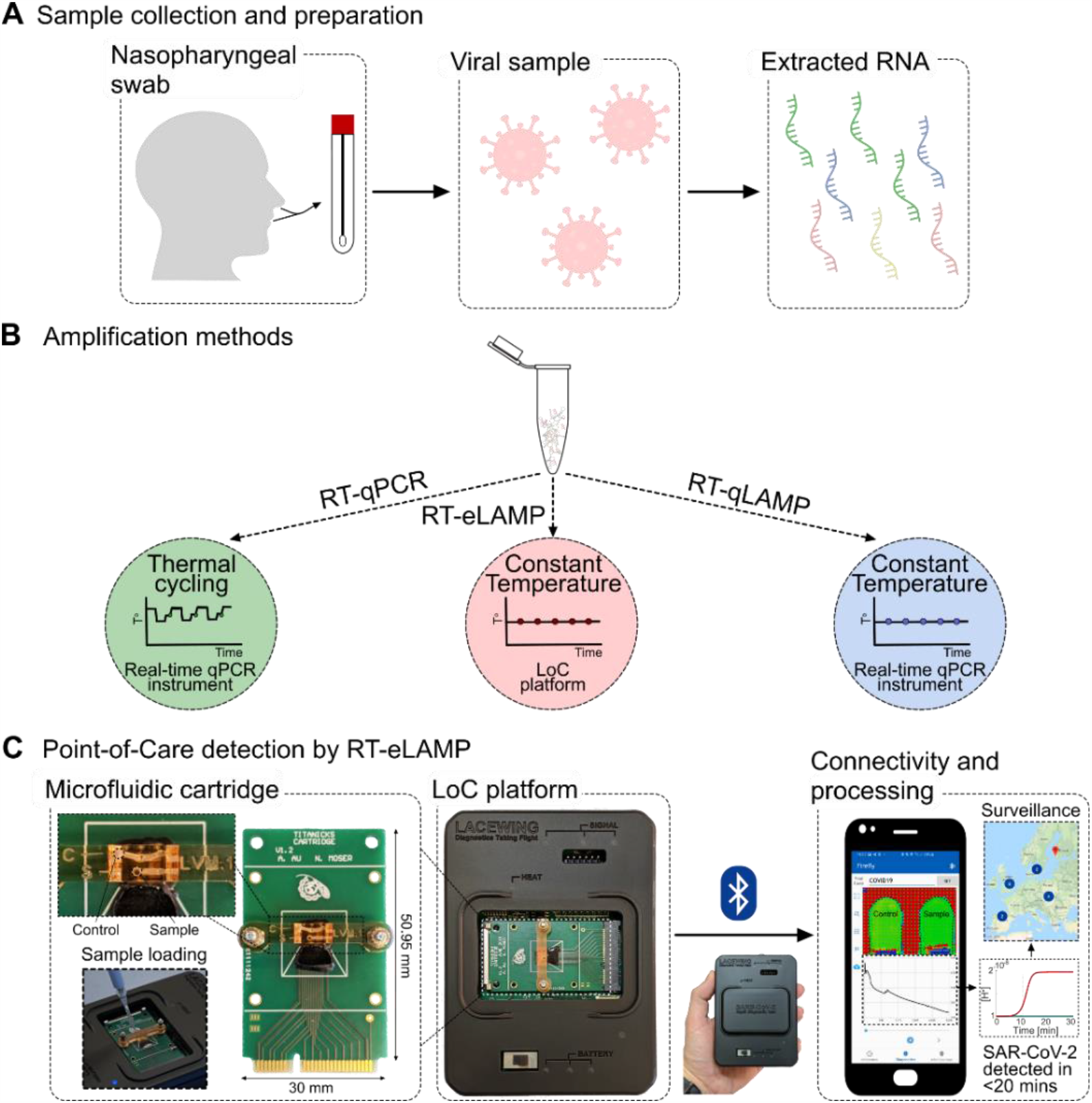
SARS-CoV-2 diagnostics workflow. **(A)** Sample collection and preparation illustrating nasopharyngeal swab and RNA extraction. **(B)** Nucleic acid amplification methods for SARS-CoV-2 RNA detection used in this study (RT-qPCR, RT-qLAMP and RT-eLAMP). Thermal profiles are illustrated for assays comparison. **(C)** Point-of-care diagnostics by RT-eLAMP showing the proposed handheld LoC platform including the microfluidic cartridge with control and sample inlets, and the smartphone-enabled application for geo-localization and real-time visualization of results.

## Results

### Optimization of LAMP assay and phylogenetic analysis for RNA detection of SARS-CoV-2

We have designed and optimized a LAMP assay targeting the N gene of SARS-CoV-2, named LAMPcov, based on previously reported LAMP assays summarized in Table S1. The N gene was selected as the optimal target since it is conserved across available sequences. The LAMP assay reported by *Zhang et al. (10)* was used as reference in this study. The LAMPcov assay was designed and optimized manually based on phylogenetic analysis of SARS-CoV-2 sequences and other related viruses (Fig. 2B). Primer sequences and location in the gene can be found in Fig. 2A and Fig. 2C. Viral sequences were retrieved from the NCBI database using nucleotide BLAST (BLASTn suite) with the amplicon of the LAMPcov assay as query. Additionally, complete genomic sequences specific to SARS-CoV-2 were retrieved from GISAID EpiCov™. A total of 8,921 sequences were collated from both databases, including sequences from different countries such as China, USA and United Kingdom. Alignments of all retrieved SARS-CoV-2 sequences were performed to evaluate *in silico* the coverage of the developed LAMP assay. Priming regions were mapped in the alignment and mismatches were found in only 195 sequences out of 8,866. These sequences differ from the reference sequence by a maximum of 2 nucleotides which indicates that our target region is specific for SARS-CoV-2 detection (Fig. S1 and S2). Furthermore, to evaluate the absence of cross-reactivity with other related viruses, a tree was built based on phylogenetic analysis of unique sequences (n = 55) using as reference the sequence NC_045512 (Fig. 2B). A clade including SARS-CoV-2, bat and pangolin coronaviruses is visible in the lower part of the tree. As previously described *(4, 26)*, bat coronavirus sequences (bat-SL-CoVZC45, bat-SL-CoVZXC21) are highly similar to the human strain. Most investigations based on genomic sequencing point towards bats and pangolins as the origin of SARS-CoV-2 with evolutionary pathways still to be established *(4)*. Therefore, non-human strains included in this clade are not anticipated to impact the outcome of the diagnostic results. Other human-infective viruses, including hCoVs such as SARS or MERS, form clades that are located at significantly higher distances. We demonstrate *in silico* the absence of cross-reactivity with other hCoVs and other related viruses based on the high number of mismatches present within the amplicon region of the proposed assay (Fig. S3). These sequences present an identity percentage below 91.4 % within the BLAST hits.

**Fig. 2:**
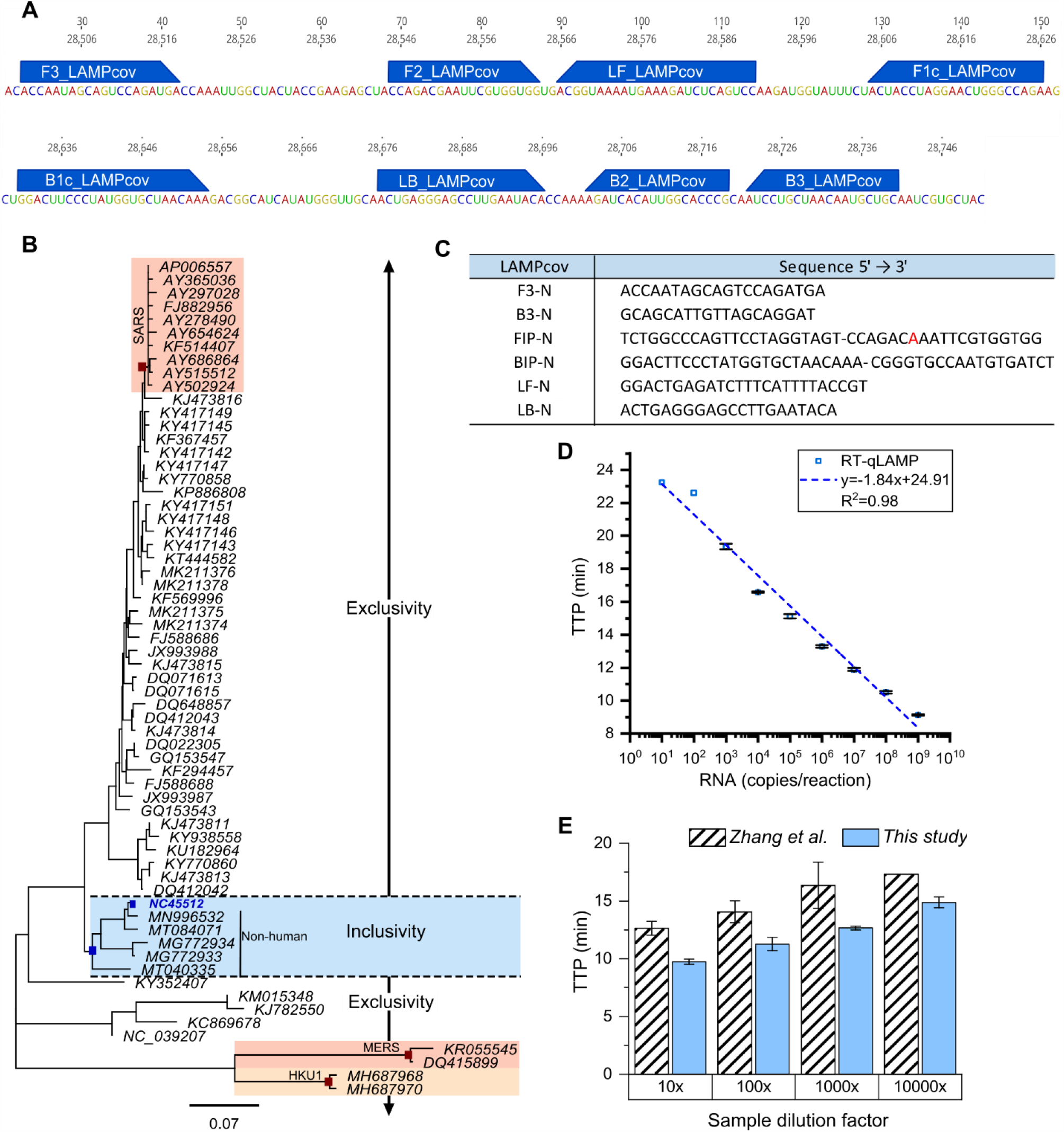
Phylogenetic analysis and LAMPcov assay design. **(A)** Reference sequence NC_45512 SARS-CoV-2 showing priming regions. **(B)** Phylogenetic tree showing the specificity of the amplicon for SARS-CoV-2 detection. Clades shadowed in blue include the reference to SARS-CoV-2 RNA sequence NC_45512. Clades highlighted in light red include HKU1, SARS and MERS, all distant from the inclusivity clade. **(C)** Sequences of primers of LAMPcov assay. One mismatch was introduced in F2 to avoid hairpin formation of the primer (in red). **(D)** Standard curve with RT-qLAMP using a control RNA at concentrations ranging between 10^1^ and 10^9^ copies per reaction. **(E)** Comparison between our assay (blue bars) and published assay by *Zhang et al. (10)* (stripped bars). Concentrations (dilution factor) of a clinical sample are plotted against TTP (min).

Analytical sensitivity of the proposed RT-qLAMP assay is shown in Fig. 2D. Time-to-positive (TTP) values ranged between 9 and 24 min, with a limit of detection of 10 copies per reaction. A standard was built (*y =* −1.84*x* + 24.91) showing a correlation of *R*^2^ *=* 0.98. The LAMP assay developed by *Zhang et al. (10)* is included as reference in Fig. 2E. The speed of our assay was increased by an average of 20% with a comparable sensitivity. Overall, results presented in Fig. 2 indicate that the proposed LAMPcov assay will not cross-react with other hCoVs or other related viruses and will be able to detect SARS-CoV-2 with high analytical specificity and sensitivity.

### Performance comparison of RT-qPCR and RT-qLAMP with clinical samples

A total of 183 clinical samples obtained from nasopharyngeal, throat or nose swabs were collected from a West London Hospital Laboratory (United Kingdom). Samples were tested with the developed LAMPcov assay and two of the PCR-based assays recommended by the CDC, the N and the RNase P assays *(7, 9)*. Calculated concentrations based on RT-qPCR and RT-qLAMP standards are detailed in Table S2. RT-qPCR results with the N assay were used as reference to classify the samples into five categories (high, upper-medium, lower-medium, low and negative) based on the *C*_*t*_ values obtained (Table 1). *C*_*t*_ was determined by the cycle-threshold method using 0.2 as the fluorescence cut-off value. This classification considered 6 cycles difference per category, which is equivalent to approximately 100-fold dilution. Results obtained with RT-qLAMP and RT-qPCR are summarized in Table 2. Sensitivity and specificity were 91% and 100% for RT-qLAMP respectively. From the 183 samples, both RT-qPCR and RT-qLAMP detected 54 samples as negative and 115 as positive. A total of 12 samples were detected as positive only by RT-qPCR (negative by RT-qLAMP) and 2 were detected as positive only by RT-qLAMP (negative by RT-qPCR). The 14 misclassified samples belong to the “Low” concentration group which could justify the discrepancy between the 2 assays. Furthermore, the 2 samples detected only by RT-qLAMP were confirmed as true positive by melting curve analysis (Fig. S4), therefore they were not considered false negative and were omitted from the sensitivity and specificity calculations (Table 2).

**Table 1:** Results obtained with RT-qPCR and RT-qLAMP.

| Cat.<sup>a</sup> | RT-qPCR C<sub>t</sub> range (cycles) | RNA conc.<sup>b</sup> (copies/μL) | Samples (n) | RT-qPCR | RT-qLAMP |
| --- | --- | --- | --- | --- | --- |
|  |  |  |  | C<sub>t</sub> ± STD (cycles) | C<sub>t</sub> ± STD (min) |
| **H** | 17 - 22.9 | 2x10<sup>6</sup> – 3x10<sup>4</sup> | 13 | 20.20 ± 1.82 | 10.08 ± 0.83 |
| **UM** | 23 - 28.9 | 3x10<sup>4</sup> – 5x10<sup>2</sup> | 36 | 26.51 ± 1.54 | 12.55 ± 1.41 |
| **LM** | 29 - 34.9 | 5x10<sup>2</sup> - 8 | 52 | 31.78 ± 1.69 | 16.47 ± 3.97 |
| **L** | > 35 | < 8 | 26 | 36.51 ± 0.88 | 21.64 ± 4.17 |
| **NEG** | NEG | / | 56 | / | / |
| **TOTAL** |  |  | 183 |  |  |

<sup>a</sup>Results are classified into categories (Cat.) based on RT-qPCR C<sub>t</sub> values. H: High, UM: upper-medium, LM: lower-medium, L: low, NEG: negative.
<sup>b</sup>RNA estimated concentration (copies/μL of eluted sample volume) based on RT-qPCR standard curve $y = -3.30x + 37.98$ .

**Table 2:** Sensitivity and specificity of RT-qLAMP compared to RT-qPCR for SARS-CoV-2 detection.

| Assay | RT-qPCR Positive (n) | RT-qPCR Negative (n) | Sensitivity<sup>a</sup> (%) | Specificity<sup>b</sup> (%) |
| --- | --- | --- | --- | --- |
| **RT-qLAMP Positive (n)** | **115** | **0\*** | 90.55 | 100 |
| **RT-qLAMP Negative (n)** | 12 | **54** |  |  |

\*Melting curve analysis (Fig. S4) confirmed specificity of the product for 2 samples detected only by RT-qLAMP, omitted in the table for sensitivity and specificity analysis.
<sup>a</sup>Sensitivity calculated based on: $SEN (\%) = \frac{TP}{TP+FN} \times 100$
<sup>b</sup>Specificity calculated based on: $SPE (\%) = \frac{TN}{TN+FP} \times 100$

To further evaluate the performance of the proposed RT-qLAMP assay in comparison to RT-qPCR, correlation between the two methods was computed by applying linear regression. A total of 115 samples were detected by both methods and included for this analysis. Viral load was calculated based on RT-qPCR and RT-qLAMP standards previously built with a control RNA (obtained from SARS-CoV-2 viral culture). Results are summarized in Fig. 3A where calculated viral loads with each of the methods were plotted. The x-axis corresponds to viral load by RT-qPCR and y-axis corresponds to viral load by RT-qLAMP. The obtained correlation is 0.8 with an *R*^*2*^ *=* 0.7*2* (n = 115). Furthermore, the RNase P assay recommended by the CDC *(9)* was used to evaluate the quality of the extracted RNA and verify the human-origin of the clinical samples. RNase P concentration was calculated based on an RT-qPCR standard previously built with a positive control provided by IDT (Hs_RPP30). The homogeneous distribution of the RNase P concentration across RT-qPCR positive (n = 127) samples for SARS-CoV-2 indicates that all samples present a similar quality after RNA extraction (Fig. 3B). However, distribution of RNase P concentration is significantly different (p-value=8.09×10^−6^) between RT-qPCR positive (n = 127) and negative (n = 56) samples for SARS-CoV-2 (Fig. 3C). This indicates that the quality of RNA might be different between both groups which could lead to an increased number of false negatives.

**Fig. 3:**
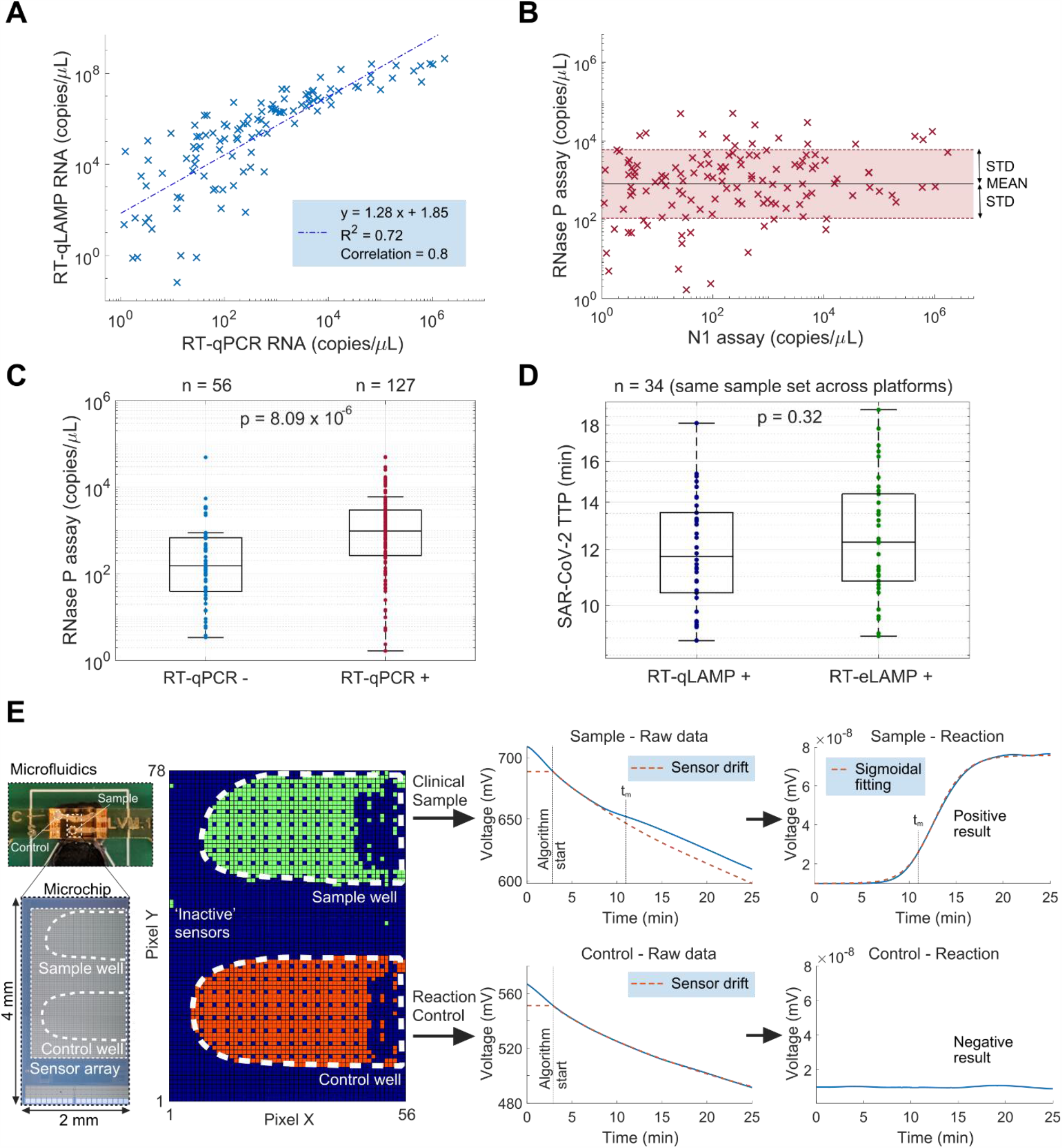
Clinical validation using RT-qPCR, RT-qLAMP and RT-eLAMP. **(A)** Correlation between RT-qPCR and RT-qLAMP based on the estimated sample concentration (copies/μL of eluted sample volume), n = 115. **(B)** Correlation between RNase P and viral RNA concentration of the clinical samples (copies/µL of eluted sample volume) to indicate quality of extraction across all the cohort, n = 127. **(C)** Boxplot distribution of RNase P concentration (copies/µL of eluted sample volume) across negative (n = 56) and positive (n = 127) clinical samples by RT-qPCR. Calculated p-value between both groups was below 0.05 (p-value = 8.09×10^−6^). **(D)** Boxplot distribution of TTP for the RT-qLAMP and RT-eLAMP demonstrating the performance of the LoC platform. qPCR. Calculated p-value between both groups is higher than 0.05 (p-value = 0.32). **(E)** Example overview of data processing steps for sample #179 to extract amplification curves from sample and control wells on the LoC platform during RT-eLAMP. The spatial image illustrates the microchip ISFET sensing array output (4,368 sensors, 2 × 4 mm) on the single-use cartridge where the averaged sensor signals in each well demonstrate respectively amplification with a TTP of 10.63 min (sample) and no amplification (control).

Overall, despite the lower sensitivity of the RT-qLAMP assay, it showed higher specificity as it detected 2 samples which were missed by the RT-qPCR assay. The RT-qPCR assay also required an additional 40 min compared to RT-qLAMP for sample screening, as at least 60 min are needed to complete the 45 recommended cycles. In particular, the average TTP values within the “High” concentration group are 20.20 ± 1.82 cycles and 10.08 ± 0.83 min, with RT-qPCR and RT-qLAMP respectively; and the average TTP values of the samples within the “Low” concentration group are 36.51 ± 0.88 cycles and 21.64 ± 4.17 min, respectively. Furthermore, RT-qPCR relies on thermal cycling, which limits its implementation for SARS-CoV-2 diagnostics at the PoC.

### Rapid point-of-care detection of SARS-CoV-2 using a Lab-on-Chip device

A subset of 40 samples (34 positive and 6 negative) were tested by RT-eLAMP to characterize the performance of our PoC test compared to a benchtop instrument. The sample size was calculated based on the equation reported by *Banoo et al. (27)*. TTP values observed for each sample are included in Table S3. The results shown in Fig. 3D demonstrate good correlation between the TTP obtained by RT-qLAMP and RT-eLAMP, achieving respectively average TTP values of 12.10 ± 2.20 min and 12.68 ± 2.56 min. Calculated p-value is above 0.05 (p-value = 0.32) demonstrating that there is no statistical difference in the performance of both platforms. This validates the incorporation of our new RT-qLAMP assay with our LoC platform.

The single-use cartridge consists of a CMOS ISFET microchip (array of 78×56 sensors, 2 × 4 mm) with a bespoke microfluidic module to accommodate two wells on the sensing surface, for sample and control reactions. An initial calibration phase relying on machine learning and spatial clustering techniques was run on the 4,368 time series from the sensor outputs to filter out sensors which were not in contact with the solution or out of readout range, labelled ‘inactive’ sensors (shown in dark blue in Fig. 3E). The remaining sensors match the positions of the sample and control wells. The algorithm averaged the output of the sensors within each well and performed drift compensation. To accurately identify the voltage change as a result of pH variations, we relied on: (i) identifying the time of maximum derivative, and (ii) performing compensation of sensor drift based on exponential interpolation to extract information on nucleic acid amplification *(23, 24, 28)*. The algorithm produced a standard amplification curve for each well, monitoring the proton concentration in real time during the reaction and hence the increase in DNA copies. Sigmoidal fitting was performed on the compensated sensor signal for detected amplifications to reflect the dynamics of nucleic acid amplification. In each experiment, we ensured that no amplification occurred in the control well within 20 min and quantified the presence of SARS-CoV-2 RNA in the clinical sample well by extracting the TTP based on 0.2 thresholding. Estimated concentration with RT-eLAMP are provided in Table S2.

The integrated sensors and novel algorithms allow to detect and quantify the presence of SARS-CoV-2 in the extracted clinical samples, with similar performance between the RT-qLAMP and the RT-eLAMP. Adding to the benefits of speed and portability, the LoC platform was connected to a custom smartphone application compatible with any Android device, and designed to acquire and process the sensor data during the reaction (Fig. 1C). Upon termination of the reaction, the data was synchronized to a secure Amazon Cloud Server (AWS) and the GPS location from the phone was used to locate the experiment on a map.

## Discussion

Stringent mitigation measures have been implemented in an effort to limit the spread of SARS-CoV-2. With 7.8 million reported cases *(1)*, the COVID-19 pandemic has been deemed a global health emergency, characterized by the uncontrolled rate of person to person transmission and ongoing increase of cases *(29, 30)*. Given the absence of a generic antiviral treatment or targeted vaccine, rapid PoC diagnostics are more critical than ever, helping to identify and track the spread of the disease.

In this paper, we combined LAMP with an *in-house* LoC device to develop a rapid PoC diagnostic test (< 20 min) for the detection of SARS-CoV-2 RNA from extracted samples. The proposed LAMP assay, named LAMPcov, was designed based on phylogenetic analysis of 8,921 sequences retrieved from NCBI and GISAID databases *(25, 31)*. The N gene was selected as the target due to its high specificity for SARS-CoV-2 detection and high conservation amongst reported SARS-CoV-2 sequences from different countries including United Kingdom, China and USA (Fig. 2). A total of 183 clinical samples were screened by RT-qLAMP and RT-qPCR using the kit recommended from the CDC (N and RNase P assays) *(9)*. Currently, RT-qPCR is considered the gold standard for SARS-CoV-2 detection. However, this method is hindered by the need for laboratory-based equipment, trained personnel, thermal-cycling instruments and procurement delays in public health institutions such as hospitals or diagnostic centers where availability of these tests is restricted to. Although the RT-qLAMP assay showed a 91% sensitivity compared to RT-qPCR, it demonstrated higher specificity since 2 samples were only detected by RT-qLAMP (specificity of the reaction demonstrated by melting curve analysis). It is important to notice that RNase P analysis showed a significant difference between positive and negative samples (p-value < 0.05) which could lead to an increase in the number of false negatives. RT-qLAMP also showed faster turnaround time with an average TTP of 15.45 ± 4.43 min. Therefore, the proposed assay is an ideal alternative to RT-qPCR for SARS-CoV-2 PoC diagnostic applications. The LAMPcov assay was also successfully implemented on the *in-house* developed LoC device and a subset of samples (n=40) were screened. Results obtained on the LoC platform presented good correlation compared to the experiments carried out in a commercial PCR thermal-cycling instrument (p-value > 0.05), confirming the robustness of the assay and the device. Currently, very few portable instruments are available for PoC diagnostics. Most of the reported rapid diagnostic tests rely on antigen or antibody detection. Antigen detection tests commonly suffer from low sensitivity, with high false negative rates reported. Nevertheless, they are potential candidates as first line rapid tests if sensitivity is improved. Antibody tests, more commonly known as serological tests, rely on the detection of antibodies such as IgA, IgM and IgG which are the product of the immune system response to SARS-CoV-2. However, several days/weeks are needed for the antibodies to be released by the immune system, shifting the detection window away from the onset of symptomatic illness. Unfortunately, it is during this window that isolation is most critical. Furthermore, antibodies are often not SARS-CoV-2 specific, leading to a high false positive rate *(6, 32)*.

Latest PoC diagnostic tests based on nucleic acid amplification methods that have been authorized by the FDA^1^ include: (1) Abbot ID NOW COVID-19 (Abbot), (2) Xpert® Xpress SARS-CoV-2 (Cepheid) and (3) AcculaSARS-Cov-2 Test (Mesa Biotech Inc.) *(33)*. The first two are quantitative tests and the last one is qualitative. All of them have an integrated RNA extraction step and provide sample-to-results within 15 to 45 min. Although most details are not disclosed, these instruments are not portable and may not be affordable for most clinics in resource-limited settings. Notably, the rapid PoC diagnostic test proposed in this work is portable and affordable due to its use of CMOS-based technology. In contrast to a recently reported lateral flow test based on CRISPR–Cas12 *(29)*, the presented device is also a quantitative test, providing information on the viral load, which is an important indicator of the patient’s infectivity. Furthermore, the proposed PoC diagnostic test can be rapidly adapted for the detection of other respiratory pathogens or infectious diseases *(22–24)* as only a target-specific LAMP assay has to be incorporated. This inherent capability promotes versatility and improves scalability, offering a wide range of applications that can be easily integrated for the simultaneous multiple detection of pathogens. This versatility further improves when considering that it can be integrated as a two-step approach with sample preparation methods or modules to obtain a sample-to-result diagnostic test which can be used for a variety of targets, from any sample type and without the need for trained personnel*(34)*. This highlights the opportunity to consider this device for use in the community, where it has the potential to offer ease of use and rapid detection through a custom smartphone app. Given the additional benefit of the incorporated geo-localization feature, we envision this PoC device as a valuable tool for epidemiological surveillance and infection tracking not only for SARS-CoV-2 but also other infectious diseases.

## Materials and Methods

### Study design

The goal of this study was to demonstrate a PoC diagnostic test based on LAMP and semiconductor technology to detect the presence of SARS-CoV-2 from extracted RNA. To evaluate performance of the proposed diagnostic test compared to reference methods, a total of 183 extracted RNA samples from nasopharyngeal, throat and nose swabs were screened by RT-qLAMP and RT-qPCR (CDC assay) *(9)*. All studies were conducted in accordance with the ethical approval (20/HRA/1561; DOCUMAS 20SM5875).

To estimate the number of samples required to be screened on the LoC platform the following formula was used *(27)*:

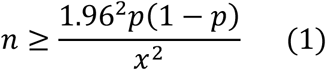

where *p* is the suspected sensitivity and *x* is the desired margin of error. We define the true-positive rate (sensitivity) as the proportion of SARS-CoV-2 positive which are correctly identified by the LoC platform compared to the benchtop instrument. We suspected the sensitivity and specificity to be 90%, similar to the one obtained with RT-qLAMP, with a desired margin of error of 10%. Under these conditions, the number of required samples is 34.57 (rounded up to 35) therefore we tested a total 40 samples (34 positive and 6 negative).

### Phylogenetics analysis

Genomic viral sequences were retrieved from NCBI using nucleotide BLAST with the amplicon of the proposed RT-LAMP assay as query (n = 4506). A total of 4451 sequences were specific for SARS-CoV-2 and 55 sequences were related to other viruses. Complete genomic sequences specific to SARS-CoV-2 were collated from GISAID EpiCov™ database using as query human host and different locations including United Kingdom, China, and USA (n = 4415). Nucleotide sequence alignment was performed using MUSCLE algorithm *(35)* in Geneious® Prime software *(36)* v2019.04 using as reference the sequence NC_045512. Neighbor-joining method was used to build a phylogenetic tree with 100 bootstraps replicates using the alignment of unique sequences retrieved from the Blastn query in NCBI (n = 55) with NC_045512 as reference for SARS-CoV-2. Accession numbers are listed in Fig. S1, S2 and S3.

### LAMP assay development

Primers were designed and optimized manually based on *Zhang et al. (10)* assay and phylogenetics analysis of SARS-CoV-2 sequences and other related viruses. Developed primers were analyzed *in silico* for presence of hairpins, dimers and cross-dimers using NUPACK *(37)*, IDT OligoAnalyzer software and ThermoFisher Multiple Primer analyzer tool. Melting temperatures (*T*_*m*_) were calculated using J. SantaLucia thermodynamics table *(38)*. All primers were synthesized by IDT and sequences are listed in Fig. 2.

### Clinical samples and controls

Clinical nasopharyngeal, throat and nose swabs were collected and processed by the Microbiology laboratory at a West London Hospital Laboratory (United Kingdom). A total of 183 samples were collected and RNA extracted using AusDiagnostics MT extractor with an elution volume ranging from 50 µL to 200 µL (AusDiagnostics) following manufacturer instructions.

Quantified RNA from viral culture was used as control. VeroE6 cells were infected with A/SARS-CoV-2/England/IC19/2020 at a MOI of 0.01 and incubated in 5% CO2 at 37 °C. After 1 hour of virus adsorption, the virus inoculum was removed and the cells were washed in phosphate-buffered saline (PBS) and replenished with Dulbecco’s Modified Eagle’s Medium (DMEM) (Gibco, Thermo Fisher Scientific) supplemented with 1% penicillin-streptomycin (Thermo Fisher Scientific) and 1% MEM non-essential amino acids (Gibco,Thermo Fisher Scientific) in 5% CO2 at 37 °C. 48 hours later, the supernatant was removed and total RNA was extracted using TRIzol reagent (Invitrogen) followed by column purification using the RNeasy RNA Mini kit (Qiagen).

### Reaction conditions

#### RT-qPCR reaction

Experiments were carried out in duplicates with a final volume of 10 μL per reaction using two of the assays recommended by the CDC, the N assay (N1 primer mix) and the RNase P assay, with the GoTaq® 1-Step RT-qPCR kit (Promega). Each mix contained the following: 10 μL of 2× GoTaq® qPCR master mix, 0.4 μL of 50× GoScript™ RT mix for 1-Step RT-qPCR, 1.2 μL of 16.6× N1/RNase P assay primer mix by the CDC (Forward Primer (20 μM), Reverse Primer (20 μM), Probe (5 μM)), 4 μL of extracted RNA and enough nuclease-free water (GoTaq® 1-Step RT-qPCR kit, Promega) to bring the volume to 20 μL. Reactions were performed following the recommendations of the manufacturer: 15 min at 63 °C for cDNA conversion, 2 min at 95 °C for AMV deactivation, and 45 cycles at 95 °C for 15 s and 55 °C for 45 s. Reactions were plated in 96-well plates and loaded into a LightCycler 96 Real-Time PCR System (LC96) (Roche Diagnostics). The N1 assay and the RNase P assay were purchased from IDT and used following the recommendations of the manufacturer.

#### RT-qLAMP reaction

Experiments were carried out in duplicates with a final volume of 10 μL per reaction. Each mix contained the following: 2 μL of 10× isothermal buffer (New England Biolabs), 1.20 μL of MgSO4 (100 mM stock), 2.80 μL of dNTPs (10 mM stock), 0.50 μL of BSA (20 mg/mL stock), 3.20 μL of Betaine (5M stock), 0.50 μL of SYTO 9 Green (20 μM stock), 1.25 μL of Bst 2.0 DNA polymerase (8,000 U/mL stock), 0.64 μL of AMV (25 U/ μL stock, Promega), 0.20 μL of Rnase inhibitor (20 U/μL stock, ThermoFisher Scientific), 4 μL of extracted RNA, 1 μL of 10 × LAMP primer mixture (20 μM of BIP/FIP, 10 μM of LF/LB, and 2.5 μM B3/F3) and enough nuclease-free water (ThermoFisher Scientific) to bring the volume to 20 μL. Reactions were performed at 63 °C for 30 min. One melting cycle was performed at 0.1 °C/s from 65 °C up to 97 °C for validation of the specificity of the products. Reactions were plated in 96-well plates and loaded into a LightCycler 96 Real-Time PCR System (LC96) (Roche Diagnostics).

#### RT-eLAMP reaction

Experiments were carried out at a final volume of 5 μL per reaction. Each mix contained the following: 0.50 μL of 10 × isothermal pH-based buffer (pH 8.5–9), 0.30 μL of MgSO4 (100 mM stock), 0.28 μL of dNTPs (25 mM stock), 0.30 μL of BSA (20 mg/mL stock), 0.13 μL of NaOH (200 mM stock), 0.80 μL of Betaine (5M stock), 0.02 μL of Bst 2.0 DNA polymerase (120,000 U/mL stock), 0.13 μL of AMV (25 U/μL stock, Promega), 0.05 μL of Rnase inhibitor (20 U/μL stock, ThermoFisher Scientific), 2 μL of extracted RNA and 0.50 μL of 10 × LAMP primer mixture (20 μM of BIP/FIP, 10 μM of LF/LB, and 2.5 μM B3/F3). Experiments were performed at 63 °C for 30 min. Reactions were loaded into a disposable cartridge and experiments were carried out using our *in-house* LoC platform.

#### Lab-on-Chip Platform

We translated the RT-LAMP assay on a portable Lab-on-Chip platform illustrated in Fig. 1C and achieving RT-eLAMP at the PoC. The platform relies on a single-use cartridge which combines state-of-the-art microchip technology and microfluidic developments. The RT-LAMP assay was performed on a large array of over 4,368 sensors called Ion-Sensitive Field-Effect Transistors (ISFETs) monitoring nucleic acid amplification through the release of protons in reaction associated with nucleotide incorporation during LAMP. Bolted onto each cartridge was a 3D printed microfluidic manifold, printed using the Fig. 4® Standalone 3D printer from 3D Systems, in their transparent bio-compatible capable (ISO 10993-5 & −10) MED-AMB-10 resin. Each manifold contained two internal microfluidic channels, which distributed 5 μL of sample and control reaction mix onto two separate designated sensing areas of the chip surface. Sealing between the manifold and chip surface was accomplished through laser-cut double-stick Tessa® adhesive gaskets, with an Ag/AgCl reference electrode (chloridised 0.03mm Ag wire) running through each reaction well between the chip surface and tape gasket. The adhesive gasket and printed resin materials were chosen for their low reaction inhibition, determined experimentally.

The battery-powered portable LoC device performed isothermal temperature regulation of the solution with a Peltier module in contact with the cartridge. An embedded microcontroller was used to implement the PID controller and transmit the data via Bluetooth to a custom smartphone application developed on AndroidOS. The app shared data on a secure cloud server hosted by Amazon Web Services and performed geo-tagging using the phone GPS.

Lab-on-Chip reactions were performed by injecting sample and control reaction mix onto each of the two designated wells (sample and control wells). Sample reaction mix contained the reaction mix described for RT-eLAMP including extracted RNA. Control reaction mix did not include extracted RNA but nuclease-free water instead. Reactions were performed for 30 min and data was recorded *in situ* in real-time. Data was analyzed in MATLAB by extracting the signal associated with pH change.

### Data analysis and statistics

Data analysis was performed using MATLAB (R2018b) and OriginPro 2019. All the TTP values are reported as mean ± std using built in functions in MATLAB. One-sample Kolmogorov-Smirnoff Test was performed to verify that the data is normally distributed; two-sample F-test was performed to verify that the data is normally distributed with equal variance; two-sample Student’s t-test was performed for comparison between groups which present normal distribution with equal mean and variance. A p-value *≤* 0.05 was considered as a threshold for statistical significance.

## Data Availability

All data available upon request.

## Supplementary Materials

Fig. S1. Alignment of SARS-CoV-2 sequences retrieved from NCBI.

Fig. S2. Alignment of SARS-CoV-2 sequences retrieved from GISAID.

Fig. S3. Alignment of SARS-CoV-2 related unique sequences retrieved from NCBI used to build a phylogenetic tree.

Fig. S4. Melting curves of samples only detected as positive by RT-qLAMP.

Table S1. Summary of reported assays for nucleic-acid amplification of SARS-CoV-2.

Table S2. Clinical samples collected in this study.

Table S3. Clinical samples tested on our Lab-on-Chip Platform by RT-eLAMP.

## Author contributions

P.G., J.R-M. and A.H. conceptualized and designed the study. K.M-C., N.M., I.P., M.C., L.M. and A.M. performed the experiments. K.M-C. and N.M. analyzed the data. G.S., P.R., F.D., F.B. R.P. and W.B. collected the samples and controls used in this study. J.R-M., K.M-C. and N. M. wrote the manuscript. All authors reviewed the manuscript.

## Data and materials availability

all data associated with this study are present in the paper or the Supplementary Materials.

## Footnotes

1 Authorized under an emergency use authorization for use by authorized laboratories and patient care settings.

